# Validation of the Perimenopause Symptom Scale (Peri-SS) for Digital Self-Assessment: Psychometric Evaluation Among U.S. Women Aged 35 to 59 years

**DOI:** 10.1101/2025.07.16.25331654

**Authors:** Yihan Xu, Irina Ilyich, Carley Prentice, Yella Hewings-Martin, Adam Cunningham, Chrisandra Shufelt, Stephanie Faubion, Liudmila Zhaunova

**Affiliations:** Flo Health Inc., London, UK; Division of General Internal Medicine, Mayo Clinic, Jacksonville, Florida, USA; Mayo Clinic Center for Women’s Health, Rochester, Minnesota, USA; Women’s Health Research Center, Mayo Clinic, Rochester, Minnesota, USA

**Keywords:** Perimenopause, perimenopause symptom scale (Peri-SS), psychometric evaluation, reliability, validity, digital health

## Abstract

**Objective:** Perimenopause is an under-recognized transition poorly measured by existing tools, which often overlook symptoms characteristic of this phase. To address this, we developed and validated the Perimenopause Symptom Scale (Peri-SS): a concise, phase-specific, psychometrically robust instrument for digital self-assessment and research use among U.S. women aged 35-59 who self-reported being in perimenopause.

**Methods:** We conducted a longitudinal validation study with 1,255 U.S. women aged 35-59 (633 self-identified as perimenopausal) recruited via Prolific. Participants completed the Peri-SS, Menopause Rating Scale (MRS), and EuroQol 5-Dimension, 5-Level (EQ-5D-5L) at baseline and 7-14 days later. Following COSMIN guidelines, we assessed structural validity, internal consistency, test-retest reliability, criterion and construct validity, and responsiveness to change.

**Results:** The Peri-SS demonstrated strong psychometric performance. Confirmatory factor analysis confirmed a four-domain structure (vasomotor, psychological, sexual, physical), with excellent model fit (CFI = 0.932, RMSEA = 0.069). Internal consistency was excellent (α = 0.90 overall; domain range 0.73-0.86), and test-retest reliability was high (ICC = 0.82). Criterion validity was strong, with Peri-SS scores highly correlated with MRS (r = 0.88). Construct validity was supported through moderate correlations with EQ-5D-5L utilities (ρ = -0.50) and effective discrimination between groups by age, menopausal status, and hormone therapy use. The scale was also responsive to symptom changes over time (η² = 0.277).

**Conclusions:** The Peri-SS is a reliable, valid, responsive tool for assessing perimenopausal symptom burden. Its concise format and strong psychometric performance make it well-suited for research, clinical care, and digital health applications.

**Two sentence summary:** The Perimenopause Symptom Scale (Peri-SS) is a novel 16-item instrument developed to measure symptom burden during perimenopause across four domains. In a validation study among U.S. women aged 35-59 years, the Peri-SS demonstrated strong reliability, validity, and responsiveness, supporting its use in research, clinical care, and digital health settings.

## Introduction

Perimenopause is a transitional life stage that typically begins in a woman’s 40s and often lasts 4 to 10 years before the onset of menopause, defined retrospectively after 12 consecutive months of amenorrhea ^1^. During this phase, women experience marked fluctuations of ovarian hormones, particularly erratic and declining serum concentrations of 17β-estradiol as well as changes in progesterone and follicle-stimulating hormone (FSH), which can trigger a wide range of disruptive physical, cognitive, and psychological symptoms, such as hot flashes, night sweats, disrupted sleep, mood changes, fatigue, and sexual health concerns ^2–5^. Although severity and duration vary, these symptoms can substantially impair daily functioning and quality of life ^6,7^.

Importantly, emerging research suggests that perimenopause is not simply an early or less intense version of menopause, but a distinct phase with its own symptom patterns, triggers, and psychosocial impact ^8^. For example, data from the Study of Women’s Health Across the Nation (SWAN) identified four distinct trajectories of vasomotor symptoms across the reproductive stages, many of which begin in perimenopause and exhibit patterns not seen in postmenopause ^9^. A large-scale analysis of over 145,000 digital symptom logs also found that perimenopausal women exhibit a unique pattern that is marked by the coexistence of persistent menstrual cycle-related symptoms and emerging menopause symptoms, distinct from both pre- and postmenopausal groups ^10^.

Despite being a universal and distinct experience that affects the quality of life for women, perimenopause remains poorly recognized in clinical practice and under-represented in health policy and research agendas ^11,12^. Further, many women feel underprepared for this life stage and report inadequate support from healthcare professionals and workplaces ^13,14^.

A major barrier to timely care is lack of awareness: many women are unfamiliar with the full range of symptoms that often begin in perimenopause and may mistakenly attribute them to stress, unrelated health issues, or part of “normal aging” ^13,15^. This challenge is compounded if those changes occur in the absence of classic vasomotor symptoms or menstrual cycles are still present ^1,13^, especially among women in their 30s or early 40s who may fall outside conventional expectations of menopausal age ^16^.

Another key obstacle to early recognition and better support lies in the lack of perimenopause-specific measurement tools. While several instruments assess symptoms associated with menopause, such as the Menopause Rating Scale (MRS) ^17^, the Menopause-specific Quality of Life (MENQOL) Questionnaire ^18^, the Greene Climacteric Scale ^19,20^, and the Menopause Transition Scale (MTS) ^21^, these were developed for assessment of postmenopausal women, rely on paper-based administration, and often omit emerging perimenopausal symptoms such as digestive issues and sexual health concerns ^10^.

As more women turn to digital tools to track and manage their reproductive health, including menstrual changes and perimenopausal symptoms ^22,23^, there is a growing need for validated, user-friendly instruments designed for scalable deployment in digital health settings. The MenoScale, an app-based self-assessment tool developed for the Zoe Health (a personalized nutrition and gut health science company), represents a promising step toward modernising menopause symptom tracking and self-monitoring ^24^. However, it was not specifically developed for perimenopausal populations and may not fully capture their distinct symptom experiences. Taken together, existing tools fall short in capturing the multifaceted and fluctuating symptom experience of women in perimenopause, which limits their usefulness for symptom tracking, early intervention, or integration into digital care platforms.

In response to these gaps and aligned with the research priorities in menopause ^12^, we developed and validated the Perimenopause Symptom Scale (Peri-SS): a concise, perimenopause-specific, and psychometrically robust instrument designed for use in clinical research and digital health contexts.

## Methods

### Scale development and validation study design

The development of the Peri-SS included initial item generation from literature review and clinical input, followed by item reduction through exploratory factor analysis (EFA) in a separate sample. This paper focuses primarily on validating the resulting 16-item scale using a prospective, longitudinal cohort study.

The validation procedures followed the COnsensus-based Standards for the selection of health Measurement INstruments (COSMIN) methodology for evaluating patient-reported outcome measures (PROMs) ^25^ and adhered to key reporting elements from the STROBE guidelines ^26^. The validation study was adequately powered based on the COSMIN-recommended sample size of at least 10 participants per item ^27^, and the final analytic sample exceeded these requirements. The study protocol was reviewed and approved by the Western Copernicus Group Institutional Review Board (IRB No. 20250554).

### Participants and recruitment

Participants were recruited between February 27 and March 16, 2025 via Prolific, an online platform designed specifically for research recruitment with a large standing pool of registered participants. Eligible individuals were women aged 35 to 59 years, residing in the United States, fluent in English. The surveys were administered via the survey platform SurveyMonkey. Women were excluded if they were currently pregnant, had experienced pregnancy loss within the past 90 days, had given birth in the past 12 months, had undergone a hysterectomy, or did not provide informed electronic consent (see Figure 1 for participant flow). These criteria were selected to ensure that participants were representative of the population likely to experience perimenopause symptoms, while minimizing potential confounding effects from recent pregnancy or potential surgical menopause. Participants were remunerated at a rate of $12-15 USD per hour for their time completing the surveys, and they on average spent 17 minutes in completing surveys.

**Figure 1.**
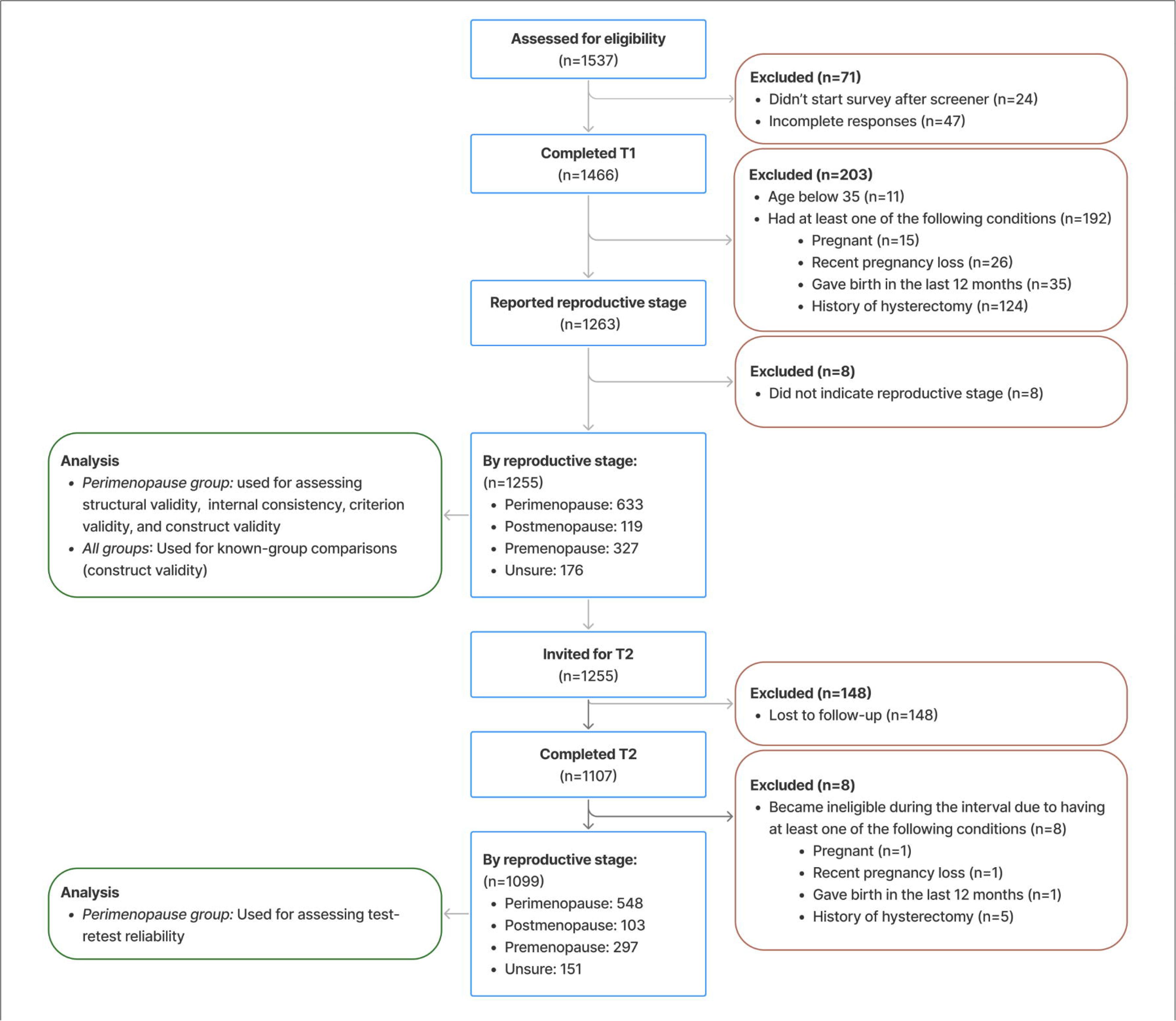
Recruitment flow.

Participants completed a baseline survey (T1) and a follow-up survey (T2), 7 to 14 days later. This interval was chosen to minimize recall bias and avoid substantial natural variation in symptom severity while allowing for an assessment of temporal reliability. Participants completed the Peri-SS, Menopause Rating Scale, and EuroQol five-dimensional questionnaire (EQ-5D-5L) at both T1 and T2, whereas they answered existing medical conditions, demographics, and other questions only at T1. The self-reported reproductive stage was assessed via a single-item question, where participants selected the stage that best described them after reviewing standardized definitions and a visual timeline of the reproductive stages (see Supplementary material A). Although STRAW+10 criteria provide the gold standard framework for reproductive ageing staging ^1^, it requires detailed and reliable longitudinal menstrual history to assess cycle characteristics, which was not feasible in this cross-sectional digital survey. Therefore, we used a simple self-reported approach to select women who self-identified as being in perimenopause, consistent with previous studies ^2,3^.

### The development of the Perimenopause Symptom Scale (Peri-SS)

The Peri-SS was developed to address the lack of concise, perimenopause-specific instruments suitable for digital self-assessment. Initial item generation drew from a targeted review of more than 15 validated scales assessing menopause and perimenopause symptoms, including the Menopause Rating Scale (MRS) ^17,28^, the Greene Climacteric Scale (GCS) ^19,20^, MENQOL ^18^, and the Women’s Health Questionnaire (WHQ) ^29^—as well as input from clinical experts.

From an initial pool of 25 items proposed by gynecologists specialized in perimenopause and menopause care, 16 were retained following exploratory factor analysis (EFA). The analysis was conducted in a sample of 948 U.S.-based women who self-reported as perimenopausal as part of the Global Perimenopause Survey (data collected from December 4, 2024 to January 6, 2025; IRB No. 20244604). Participants were users of the Flo App, a female health digital platform where women could track their menstrual cycles, pregnancy, or manage perimenopause. Item selection was based on factor loadings, internal consistency, and clinical interpretability, resulting in a four-domain structure: vasomotor symptoms (2 items), psychological and emotional stress (5 items), sexual health (3 items), and physical/somatic changes (6 items).

This scale structure was reviewed by the same gynecologists to ensure clinical validity and relevance. Each item is rated on a 5-point Likert scale ranging from 0 (none) to 4 (very severe), reflecting the impact of symptoms over the past 30 days. Domain scores are computed as the mean of items within each domain, and the overall Peri-SS score is calculated as the average of domain scores and scaled to a 0-100 range, with higher scores reflecting greater symptom burden (the final item list are detailed in Supplementary material B).

To support interpretation, Peri-SS score is categorized into four severity bands, conceptually aligned with the MRS: none to mild (0-8), mild to moderate (9-18), moderate to severe (19-35), and severe (≥36). These thresholds were empirically derived using a data-driven approach and calibrated based on their alignment with benchmark measures of symptom burden and quality of life (see Supplementary Material C for derivation procedure details).

### Validation procedures

We evaluated the following psychometric properties of the Peri-SS: structural validity, criterion validity, construct validity (including hypothesis testing and known-group comparisons), internal consistency, and test-retest reliability. Where relevant, thresholds for interpretation followed COSMIN and related psychometric conventions ^25^, and we present the COSMIN checklist in Supplementary material D. All statistical analyses were conducted using R (version 4.4.1), using the “lavaan” package ^30^ for structural equation modeling and “psych” package ^31^ for reliability analyses. All reliability and validity estimates were calculated using complete-case data from T1 survey, except for test-retest reliability and responsiveness, which used data from both T1 and T2.

#### Structural validity

Construct validity assesses whether the Peri-SS accurately measures the underlying concept of perimenopause symptoms. We conducted confirmatory factor analysis (CFA) to test whether the four-factor model identified during instrument development adequately fits the observed data. The model was estimated using full-information maximum likelihood (FIML) to account for missing data and applied standardized latent variable variances. Model fit was assessed using the Comparative Fit Index (CFI) and the Root Mean Square Error of Approximation (RMSEA). Fit was considered excellent if CFI > 0.95 or RMSEA < 0.06, and acceptable if CFI > 0.90 or RMSEA < 0.08, in line with COSMIN criteria ^25^.

#### Internal consistency

Internal consistency refers to how well the items on a scale measure the same underlying concept. We assessed it using Cronbach’s alpha (α) for the total score and each of the four domains. Values of α ≥ 0.70 were considered acceptable, ≥ 0.80 good, and ≥ 0.90 excellent. Unidimensionality within domains was verified by reviewing inter-item correlations and factor loadings from the CFA.

#### Test-retest reliability

To evaluate the temporal stability of the Peri-SS, we assessed test-retest reliability over a 7-14 day interval using data from participants who completed both T1 and T2 surveys. We calculated the intraclass correlation coefficient (ICC) using a two-way mixed-effects model with absolute agreement for the total Peri-SS score.

#### Responsiveness

Responsiveness refers to the ability of an instrument to detect meaningful change over time in the construct it intends to measure. We assessed responsiveness of the Peri-SS by examining whether changes in Peri-SS scores reflected changes in symptom severity, using the MRS as an external anchor. Responsiveness of the Peri-SS was evaluated using an anchor-based approach. Change in total MRS score between T1 and T2 surveys served as the external anchor. Participants were categorized into three groups based on MRS score change between baseline and follow-up: *improved* (≥4-point reduction), *no change* (change between –4 and +4 points), or *worsened* (≥4-point increase). We then compared Peri-SS score changes across these groups using the Kruskal-Wallis test. Effect size was quantified using eta-squared based on the H-statistic (η²[H]), with thresholds of 0.01, 0.06, and 0.14 representing small, medium, and large effects, respectively ^32^.

#### Criterion validity

Criterion validity examines whether Peri-SS scores align with an established domain-specific measure. We assessed criterion validity by comparing the Peri-SS to the MRS, a widely used instrument in clinical and research contexts to evaluate menopausal symptom burden. While the MRS is not a gold standard for perimenopause-specific symptom burden, it includes overlapping domains such as psychological, somatic, and urogenital symptoms, making it a suitable reference for perimenopausal symptom burden ^17,28^.

We computed Pearson’s correlation coefficient (r) to quantify the correlation between Peri-SS and the two established scales, and we considered the correlation to be strong if r ≥ 0.70. We also computed Cohen’s kappa (κ) to assess agreement in categorical symptom severity classifications (e.g., minimal, mild, moderate, severe). The correlation coefficient was considered strong if r or κ is ≥ 0.70 ^25^.

#### Construct validity

Construct validity examines whether the Peri-SS measures the theoretical construct it is intended to measure—namely, the perimenopause symptom burden. We assessed construct validity using two complementary approaches: convergent validity and known-group validity.

For convergent validity, we evaluated the relationship between Peri-SS total scores and the EQ-5D-5L utility, a widely used measure of general health-related quality of life ^33^. The EQ-5D-5L instrument includes five dimensions (mobility, self-care, usual activities, pain/discomfort, and anxiety/depression), each rated on five levels of severity. The responses were converted into utility values using the U.S. population-specific value set ^34^. We hypothesized that greater symptom burden, as captured by the Peri-SS, would be associated with lower EQ-5D utility values. We computed correlation using Spearman’s ρ as the distribution of EQ-5D-5L scores were highly skewed. Based on COSMIN thresholds, the convergent validity was considered strong if |r| or |ρ| ≥ 0.50; weak if they are below 0.30; moderate if they’re between 0.3 and 0.5 ^25^.

For known-group validity, we compared Peri-SS scores across subgroups expected to differ in symptom severity, including age group, and reproductive stage. As Peri-SS scores were not normally distributed, we used Kruskal-Wallis tests to assess overall group differences. Where significant main effects were detected, we conducted post hoc pairwise comparisons using Wilcoxon rank-sum tests. All *p*-values from pairwise comparisons were adjusted using the Bonferroni method to account for multiple testing. We hypothesized that symptom scores would be higher among women in their mid-40s to early 50s and among those in perimenopause or postmenopause compared with premenopausal women.

## Results

### Participant characteristics

Figure 1 outlines the participant recruitment and eligibility flow. Of 1,537 individuals screened, 1,466 completed the baseline (T1) survey (95.3% completion rate), among which 1,255 met all eligibility criteria, including providing their reproductive stage. These participants formed the primary analytic sample for all baseline psychometric analyses.

The majority self-identified as perimenopausal (50.4%), with the remainder identifying as premenopausal (26.1%), postmenopausal (9.5%), or unsure (14.0%). The perimenopause subgroup (n = 633) was used for confirmatory factor analysis and internal consistency assessments. All reproductive stage groups were included in construct validity analyses. Table 1 summarizes baseline sociodemographic and clinical characteristics of the baseline sample. Among the 1,255 eligible participants who completed the T1 survey, 1,107 completed the T2 survey (88.2% completion rate), of whom 1,099 remained eligible and were included in test-retest reliability analyses (see Figure 1 for the detailed participant flow).

**Table 1.**
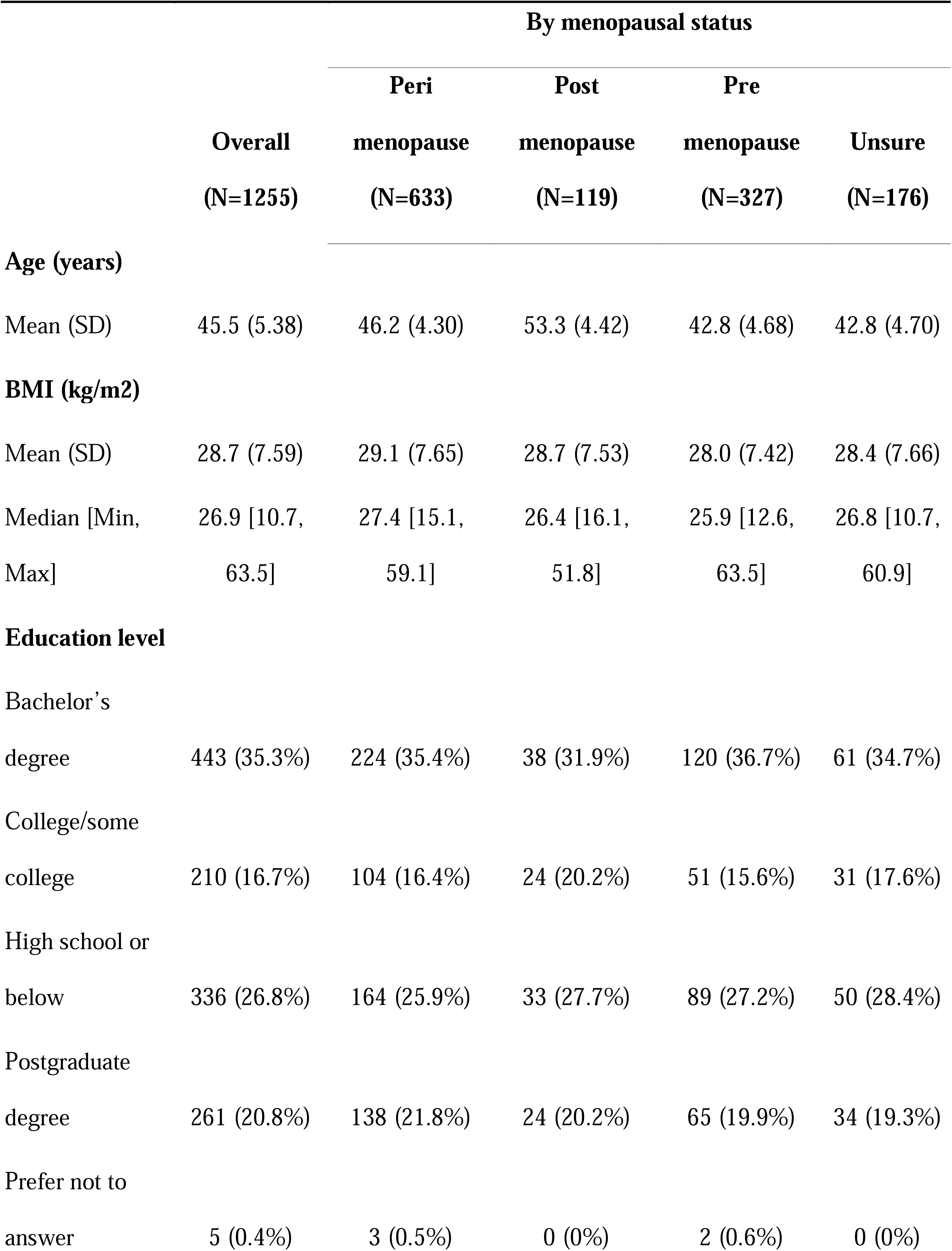
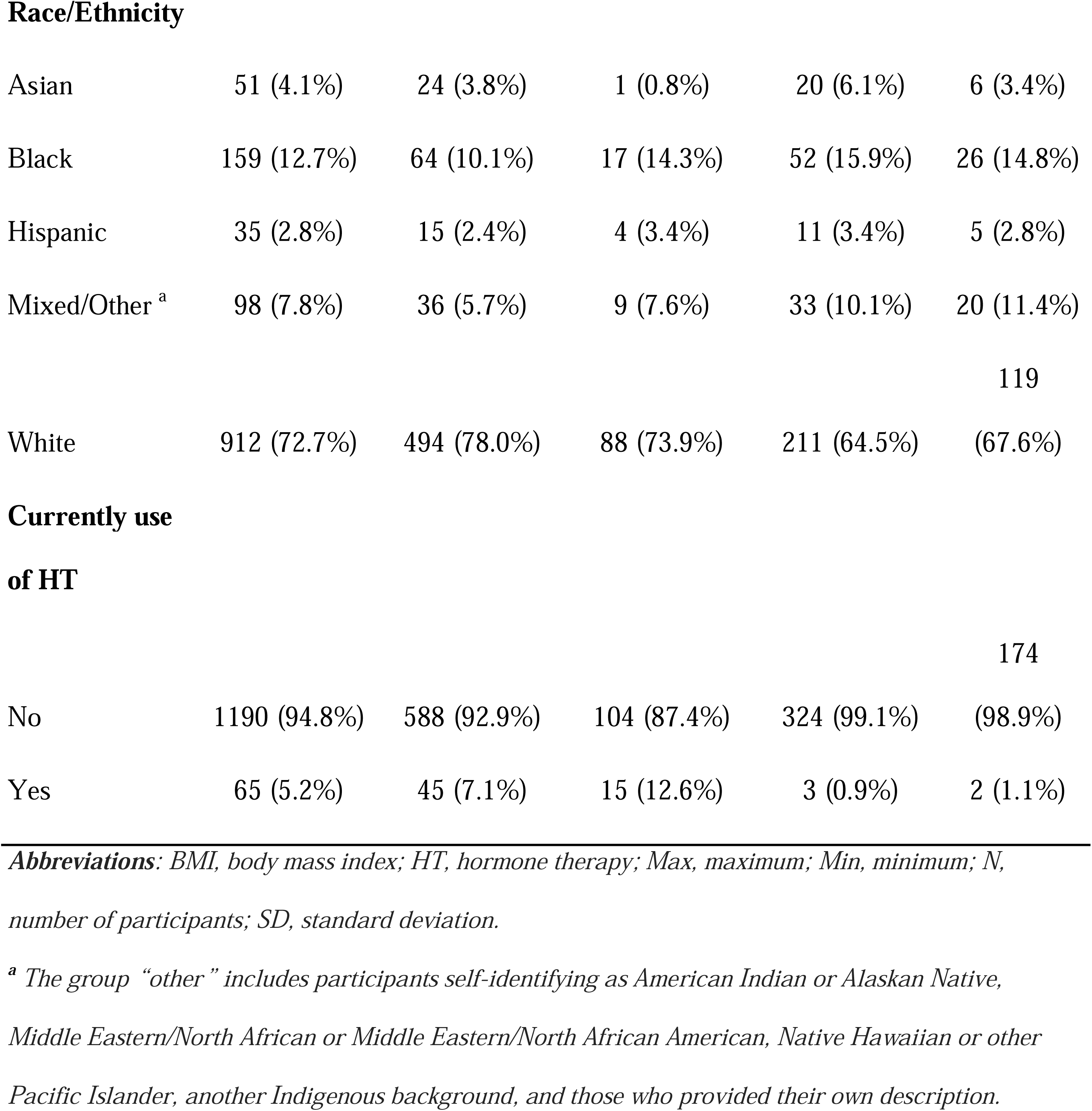
Characteristics of eligible participants at baseline.

### Structural validity

Structural validity was evaluated using confirmatory factor analysis (CFA) on the perimenopausal subgroup (n = 633) to assess whether the hypothesized four-factor model (vasomotor, psychological, sexual health, and physical / somatic domains) fit the observed data. The CFA results supported the hypothesized four-factor structure of the Peri-SS. Model fit was acceptable, with CFI = 0.932 and RMSEA = 0.069. All items loaded significantly on their intended factors, with standardized factor loadings ranging from 0.536 to 0.872 (see Table 2). These results suggest that the Peri-SS captures four distinct but correlated domains of perimenopause symptoms.

**Table 2.** Internal consistency and standardized factor loadings for the four-factor structure of the Perimenopause Symptom Scale (Peri-SS)

|  | **Vasomotor** | **Sexual** | **Psychological** | **Physical/Somatic** |
| --- | --- | --- | --- | --- |
| Cronbach's $\alpha$ | 0.73 | 0.78 | 0.86 | 0.83 |
| Hot flashes | 0.729 |  |  |  |
| Night sweats | 0.790 |  |  |  |
| Vaginal dryness |  | 0.872 |  |  |
| Pain during sex |  | 0.761 |  |  |
| Low sex drive |  | 0.604 |  |  |
| Feeling down |  |  | 0.825 |  |
| Irritability |  |  | 0.756 |  |
| Anxiety |  |  | 0.824 |  |
| Sleep difficulties |  |  | 0.555 |  |
| Physical exhaustion |  |  | 0.752 |  |
| Digestive issues |  |  |  | 0.691 |
| Hair changes |  |  |  | 0.700 |
| Skin changes |  |  |  | 0.748 |
| Joint discomfort |  |  |  | 0.654 |
| Bladder problems |  |  |  | 0.536 |
| Weight changes |  |  |  | 0.691 |

**Abbreviations:** Peri-SS, Perimenopause Symptom Scale.
**Note:** All factor loadings are standardized and statistically significant at $p < .001$ .

For comparative purposes, we also conducted CFA on the MRS. It had a slightly lower model fit, with CFI = 0.927 and RMSEA = 0.087. While all MRS items also loaded significantly on all three domains, standardized loadings were lower in the somatic and urogenital domains (e.g., 0.470-0.620 and 0.545-0.680, respectively). This comparison shows the structural coherence of the Peri-SS and its potential for capturing a broader and more differentiated range of perimenopause symptoms.

Figure 2 shows a path diagram of the final four-factor model of the Perimenopause Symptom Scale (Peri-SS) based on confirmatory factor analysis. Circles represent latent constructs (domains), and rectangles represent observed symptom items. Standardized factor loadings are shown on each path and were all statistically significant at p < .001. Double-headed arrows represent correlations between latent domains. This structure supports the theorized domains of vasomotor, sexual, psychological, and physical symptoms in the Peri-SS.

**Figure 2.**
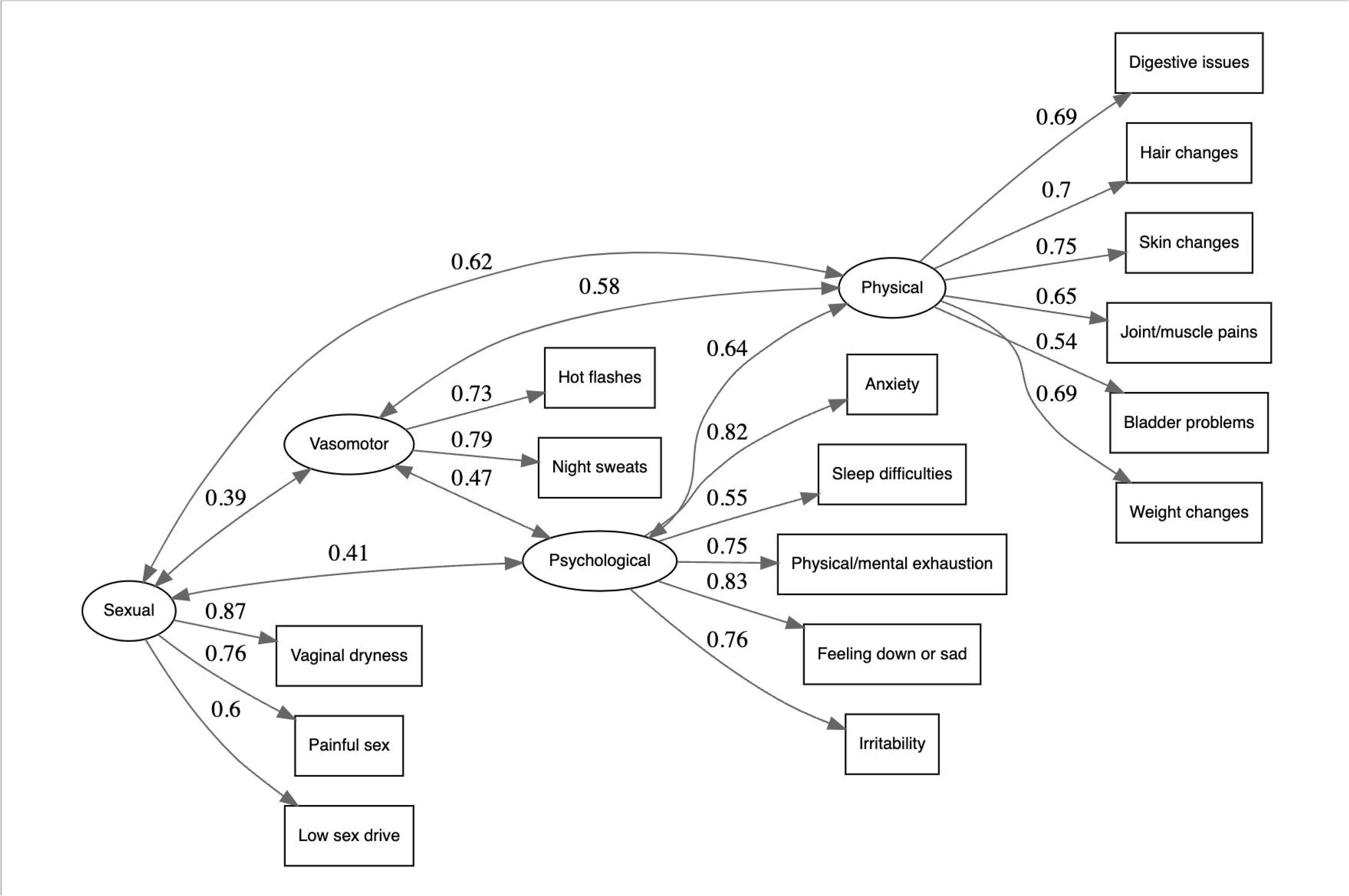
Path diagram of the final four-factor model of the Perimenopause Symptom Scale (Peri-SS) based on confirmatory factor analysis. Circles represent latent constructs (domains), and rectangles represent observed symptom items. Standardized factor loadings are shown on each path and were all statistically significant at p < .001. Double-headed arrows represent correlations between latent domains. This structure supports the theorized domains of vasomotor, sexual, psychological, and physical symptoms in the Peri-SS.

### Internal consistency

The Peri-SS demonstrated excellent overall reliability (standardized Cronbach’s α = 0.90). Subscale reliability was also strong, with α values of 0.73 for the vasomotor domain, 0.78 for the sexual domain, 0.86 for the psychological domain, and 0.83 for the physical domain. These results indicate good to excellent internal consistency across all domains, suggesting that the items within each domain reliably measure a coherent underlying construct. For comparison, the MRS showed an overall α of 0.86. Domain-specific values were 0.88 for psychological, 0.67 for somatic, and 0.62 for urogenital symptom domains. These results show that Peri-SS outperformed MRS in reliability among women who self-identified as perimenopausal.

Table 3 presents the descriptive summary of Peri-SS total score and domain scores among women self-identified as being in perimenopause, stratified by age group. The total score is scaled to range from 0 to 100, reflecting overall symptom burden, while domain scores range from 0 to 4, with higher scores indicating greater symptom severity. The average total score across all perimenopausal participants was 32.6 (SD = 16.6). Descriptively, across the four symptom domains, psychological and emotional stress emerged as the most prominent symptom domain across all age groups, while sexual health symptoms were rated the lowest. Vasomotor and sexual symptom severity generally increased with age, whereas psychological symptoms peaked in midlife and declined slightly in the oldest age group. In contrast, physical and somatic symptom scores remained relatively stable across age groups. However, Kruskal-Wallis tests found no statistically significant age-group differences in the total score or any domain score among perimenopausal participants. A more detailed breakdown of item-level scores is presented in Supplementary S-Table 2.

**Table 3.** Peri-SS total score and domain scores by age group among perimenopausal participants (N = 633)

|  | 35-44 | 45-54 | 55-59 | Overall | Kruskal- |  |
| --- | --- | --- | --- | --- | --- | --- |
| | (N=184) | (N=435) | (N=14) | (N=633) | Wallis $\chi^2$ | P value |
|  |  |  |  |  | (df) |  |
| **Total score** | 30.7 (16.6) | 33.4 (16.6) | 31.6 (14.7) | 32.6 (16.6) | 3.09 (2) | 0.213 |
| **Vasomotor** |  |  |  |  |  |  |
| **symptoms** | 1.1 (0.86) | 1.2 (0.93) | 1.3 (1.1) | 1.2 (0.91) | 1.45 (2) | 0.483 |
| **Psychological** |  |  |  |  |  |  |
| **and emotional** |  |  |  |  |  |  |
| **stress** | 1.7 (0.83) | 1.7 (0.83) | 1.2 (0.72) | 1.7 (0.83) | 5.08 (2) | 0.079 |
| **Sexual health** | 1.0 (0.94) | 1.2 (0.94) | 1.3 (1.1) | 1.1 (0.94) | 4.61 (2) | 0.1 |
| **Physical and** |  |  |  |  |  |  |
| **somatic** |  |  |  |  |  |  |
| **changes** | 1.1 (0.81) | 1.2 (0.76) | 1.2 (0.84) | 1.2 (0.78) | 5.63 (2) | 0.06 |

**Abbreviations:** *df*, degrees of freedom; *N*, number of participants; *Peri-SS*, Perimenopause Symptom Scale; *SD*, standard deviation
**Note:** Values are presented as Mean (SD). Total score ranges from 0 (minimal symptom) to 100 (severe symptom), whereas domain scores range from 0 (minimal symptom) to 4 (severe symptom). Between-group differences were assessed using the Kruskal-Wallis rank-sum test.

### Test-retest reliability

We assessed test-retest reliability among 552 participants who remained eligible and completed both T1 and T2 surveys within a 7-14 day interval. The intraclass correlation coefficient (ICC) for the total Peri-SS score was 0.82 (95% CI: 0.70-0.88), indicating excellent reliability. Domain-specific ICCs also demonstrated excellent temporal stability, ranging from 0.77 (physical), 0.80 (vasomotor), 0.80 (sexual), to 0.81 (psychological). In comparison, the total MRS score yielded an ICC of 0.79 (95% CI: 0.66-0.85), with domain-specific ICCs ranging from 0.75 (urogenital), 0.77 (psychological), to 0.77 (somatic). These results support the Peri-SS as a stable and reliable instrument for assessing perimenopause symptom burden over short intervals.

### Responsiveness

The Peri-SS was responsive to changes in symptom burden over time. As shown in Figure 3, Peri-SS score changes differed significantly across the MRS anchor categories (Kruskal-Wallis χ² = 153.1, *p* < 0.001). Participants classified as *Improved* (n = 195) showed a mean reduction of –9.5 points (SD = 8.9), while those who *Worsened* (n = 42) showed a mean increase of +8.1 points (SD = 8.7). Those with *No change* (n = 311) had a small mean decrease of –2.8 points (SD = 6.4).

**Figure 3.**
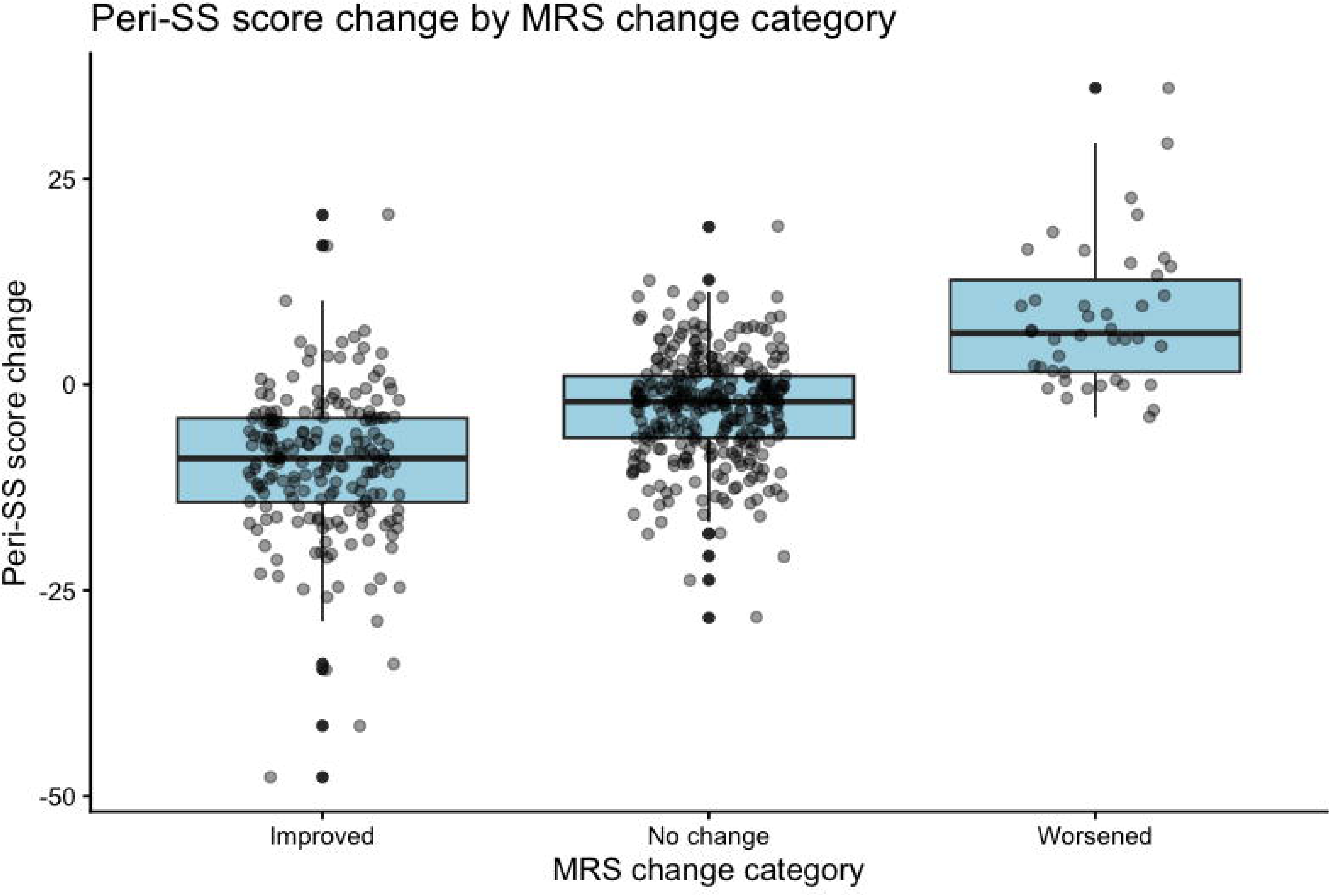
Peri-SS score change by MRS change category. Boxplots illustrate changes in Peri-SS scores from baseline to follow-up, grouped by whether participants’ symptom severity— anchored to changes in the Menopause Rating Scale (MRS)—*improved*, showed *no change*, or *worsened*. Negative values indicate symptom improvement. Peri-SS scores decreased significantly in the “improved” group and increased in the “worsened” group, supporting the scale’s responsiveness to clinically meaningful change.

The effect size for the association between MRS change category and Peri-SS score change was η² = 0.277, indicating a large magnitude of responsiveness. These results suggest that although the Peri-SS captures symptom impact over the past 30 days, it was sensitive to changes detected within a shorter interval, supporting its use for monitoring symptom progression over time.

Figure 3 presents Peri-SS score change by MRS change category. Boxplots illustrate changes in Peri-SS scores from baseline to follow-up, grouped by whether participants’ symptom severity— anchored to changes in the Menopause Rating Scale (MRS)—*improved*, showed *no change*, or *worsened*. Negative values indicate symptom improvement. Peri-SS scores decreased significantly in the “improved” group and increased in the “worsened” group, supporting the scale’s responsiveness to clinically meaningful change.

### Criterion validity

The Peri-SS showed strong criterion validity. We found a strong positive correlation between Peri-SS and MRS total scores (Pearson’s *r* = 0.88, *p* < 0.001), indicating substantial overlap in symptom severity ratings between the two instruments. A supplementary analysis using Cohen’s Kappa (Supplementary material E and S-Table 1) also showed moderate agreement between MRS and Peri-SS severity categories (κ = .58).

### Construct validity

The construct validity of the Peri-SS was supported by both convergent and known-group validity analyses. For convergent validity, the Peri-SS scores were moderately and significantly negatively correlated with EQ-5D utility values (Spearman’s ρ = – 0.50, *p* < 0.001), suggesting that greater perimenopause symptom burden was associated with poorer health-related quality of life. For comparison, the MRS demonstrated a slightly stronger correlation with EQ-5D utility values (Spearman’s ρ = –0.58, *p* < 0.001).

The Peri-SS demonstrated strong known-group validity, with significant differences observed across subgroups expected to vary in symptom burden (Figure 4, Table 4). As hypothesized, symptom scores were significantly different by age group (*χ²* = 31.4, *p* < 0.001), with women aged 45-54 years reporting higher symptom burden than those aged 35-44 (adjusted *p* < 0.001), consistent with the typical onset of perimenopause symptoms in midlife. Scores among women aged 55-59 did not significantly differ from either of the younger age groups after correction for multiple comparisons.

**Figure 4.**
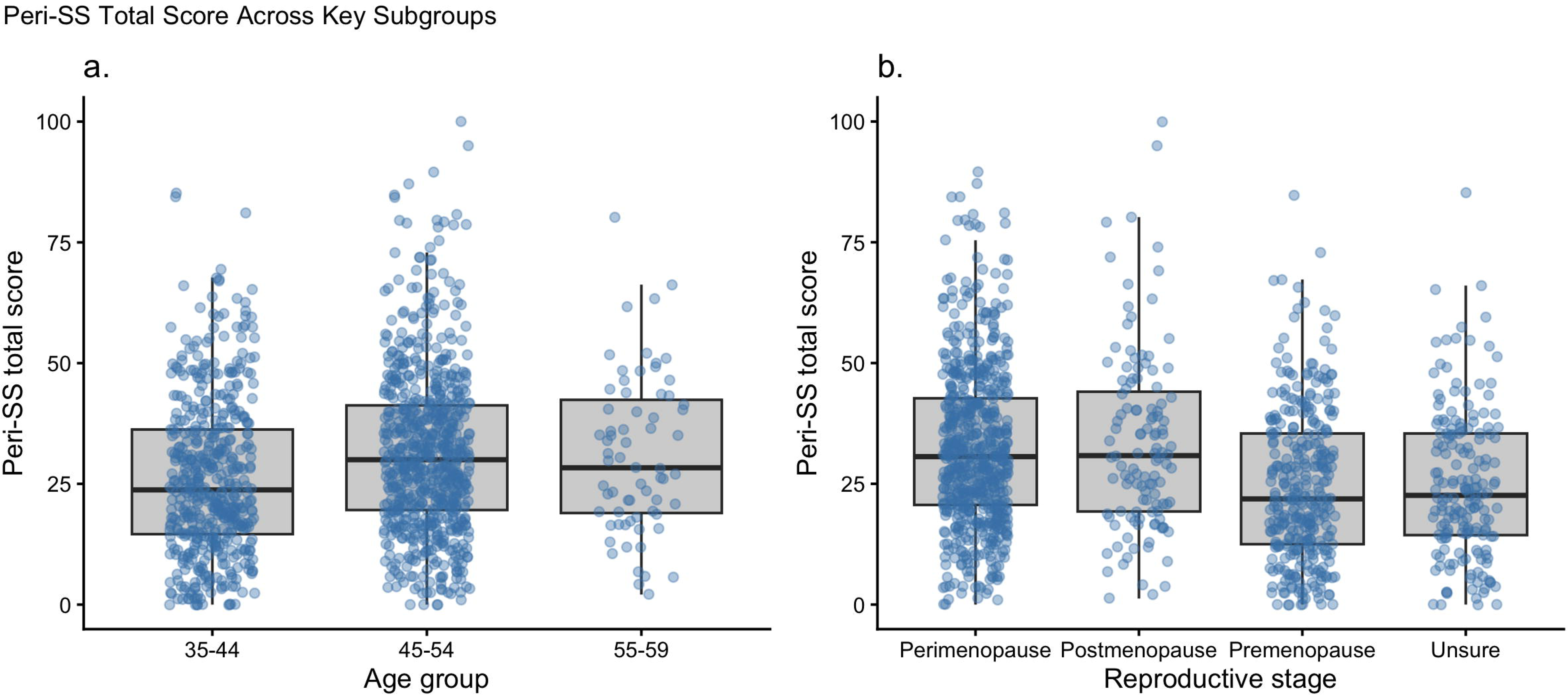
Perimenopause Symptom Scale (Peri-SS) total scores by a) age group (n = 1255) and b) reproductive stage (n = 1255). Boxplots show the median, interquartile range, and individual data points (jittered). Scores were significantly higher in the 45-54 year age group (p < .001), in the perimenopause and postmenopause group (p < .001).

**Table 4.** Descriptive statistics and known-group differences in Peri-SS total scores by age group, menopausal status, and HT use (Kruskal–Wallis Test)

| Group | Category | N | Mean (SD) | Median [Min, | Test statistic | p- |
| --- | --- | --- | --- | --- | --- | --- |
| Variable |  |  |  | Max] |  | value |
| Overall |  | 1255 | 29.5 (16.8) | 27.7 [0, 100] |  |  |
| Age group | 35–44 | 497 | 26.3 (16.2) | 23.8 [0, 85.2] | $\chi^2 = 31.4$ | <0.001 |
|  | 45–54 | 691 | 31.6 (17.0) | 30.0 [0, 100] |  |  |
|  | 55–59 | 67 | 30.9 (16.3) | 28.3 [2.1, 80.2] |  |  |
| Menopausal |  |  |  |  |  |  |
| status | Premenopause | 327 | 24.5 (15.6) | 21.9 [0, 84.8] | $\chi^2 = 71.2$ | <0.001 |
|  | Perimenopause | 633 | 32.6 (16.6) | 30.6 [0, 89.6] |  |  |
|  | Postmenopause | 119 | 33.5 (18.8) | 30.8 [1.25, 100] |  |  |
|  | Unsure | 176 | 24.8 (15.3) | 22.6 [0, 85.2] |  |  |
| HT use | No | 1190 | 29.1 (16.8) | 27.3 [0, 100] | $\chi^2 = 9.67$ | 0.018 |
|  | Yes | 65 | 35.8 (17.1) | 33.1 [3.33, 95.0] |  |  |

**Abbreviations:** HT, hormone therapy; Max, maximum; Min, minimum; N, number of participants; Peri-SS, Perimenopause Symptom Scale; SD, standard deviation.

Peri-SS scores varied significantly by reproductive stage (*χ²* = 71.2, *p* < 0.001), with perimenopausal women reporting greater symptom burden than both premenopausal and “unsure” participants (adjusted *p* < 0.001 for both comparisons). Postmenopausal women also reported significantly higher scores than premenopausal and unsure groups (adjusted *p* < 0.001), but there was no significant difference between perimenopausal and postmenopausal participants. These findings are in line with expectations that women undergoing or having recently gone through perimenopause would experience more intense symptoms than those not yet in transition or uncertain of their reproductive stage.

For comparison, we also examined known-group differences for the MRS (see Supplementary S-Table 3 and 4). While overall patterns were broadly similar, Peri-SS detected additional group differences not identified by MRS, such as between ages 35-44 years and 55-59 years, and between postmenopause and unsure groups.

Together, these results strengthen the known-group validity of the Peri-SS by demonstrating its ability to discriminate between clinically relevant subgroups, in line with theoretical expectations.

Figure 4 presents Perimenopause Symptom Scale (Peri-SS) total scores by a) age group (n = 1255), b) menopausal status (n = 1255), and c) current hormone therapy (HT) use (n = 1255). Boxplots show the median, interquartile range, and individual data points (jittered). Scores were significantly higher in the 45-54 year age group (p < .001), in the perimenopause and postmenopause group (p < .001), and in women using HT (p = .002).

## Discussion

### Principal findings

Our findings provide robust evidence that the Peri-SS, a new digital self-assessment instrument designed to measure multidimensional symptom burden in perimenopause, meets key psychometric standards. Below, we summarize the key measurement properties demonstrated through this validation study.

Structurally, the Peri-SS was confirmed to have a four-factor model (vasomotor, psychological, sexual, and physical/somatic symptoms), supported by confirmatory factor analysis with good model fit (CFI = 0.932, RMSEA = 0.069). Internal consistency was excellent, with a total Cronbach’s alpha of 0.90, and subscale alphas ranging from 0.73 to 0.86. On average, women in perimenopause reported moderate symptom burden as measured by Peri-SS, with some variation by age group. At the domain level, psychological symptoms were the most commonly reported, while sexual health symptoms were the least reported. Vasomotor and sexual symptoms showed a trend of increasing severity with age, whereas psychological symptoms appeared to peak during midlife and decline slightly in older age groups. Physical and somatic symptoms remained relatively stable across age groups.

The Peri-SS also demonstrated high test-retest reliability over a 7-14 day interval (ICC = 0.82), suggesting stable symptom reporting over short timeframes. In addition, the Peri-SS demonstrated large responsiveness to change (η² = 0.277), with symptom scores decreasing significantly among those who improved on an external anchor (MRS) and increasing among those whose symptoms worsened. In terms of criterion validity, the scale correlated strongly with the MRS (r = 0.88), supporting its ability to capture perimenopause symptom burden consistent with established instruments. Finally, construct validity was established through both convergent and known-group approaches: the Peri-SS was moderately negatively correlated with EQ-5D utility scores (ρ = –0.50), indicating that higher symptom burden was associated with lower quality of life, and was significantly associated with differences in age and reproductive stage.

Taken together, these findings provide strong initial support for the Peri-SS as a reliable, valid, and responsive instrument for assessing perimenopause symptoms among U.S. women aged 35-59 years in clinical, research and digital health settings.

### Comparison with existing instruments

To better understand the contribution of the Peri-SS, it is important to consider how it compares to existing symptom measurement tools for menopause and midlife women’s health.

A wide range of instruments exist to measure menopause symptoms, but most were developed for postmenopausal populations or general midlife health rather than the evolving needs of women in perimenopause ^35^. There remains a significant knowledge gap in effective strategies to help women prepare for and recognize the start of perimenopause ^12^. While existing instruments have proved useful, they share key limitations ^17–20,24^, validation was often done among postmenopausal women, early perimenopause symptoms were underrepresented, reliance on paper-based administration, and psychometric evaluation did not consistently adhere to best-practice psychometric standards, such as the COSMIN framework ^25^.

Among these, the MRS is the most widely validated tool and serves as the primary comparator for this study ^17^. The Peri-SS shares structural similarities with MRS—both include psychological, somatic, and urogenital symptoms—and demonstrated strong convergent validity in our analysis. Like the MRS, the Peri-SS offers clearly defined domains supported by factor-analytic results, allowing for interpretable and actionable domain-specific scores. However, the Peri-SS appears to offer improved scope and relevance for perimenopausal populations. This is because several commonly reported symptoms such as low sex drive, weight changes, and digestive issues are underrepresented in MRS. Conversely, some of its somatic items (e.g., heart palpitations) may be more characteristic of later reproductive stages and thus less reliable in perimenopause populations ^36^. Overall, Peri-SS may more accurately capture the earlier, fluctuating symptomatology typical of perimenopause, where symptom clustering may differ from that observed postmenopause ^10^.

More recently, the MenoScale ^24^ represents an important step forward in menopause symptom measurement in digital contexts. Developed using data from digital health users, it captures a broad range of symptoms across 21 items, with particular emphasis on somatic symptoms (10 items). Its factor structure, comprising vasomotor, sexual, psychological, and somatic domains, is similar to that of the Peri-SS. However, MenoScale was validated primarily among older women (average age over 50) and a demographically homogeneous sample (95% White), which may limit its applicability to younger or more diverse perimenopausal populations. In contrast, the Peri-SS was developed and validated in a younger (mean age 46), more demographically diverse cohort (73% White), and achieved robust psychometric performance across all domains while balancing symptom comprehensiveness and survey burden.

Other legacy instruments further highlight the gap Peri-SS fills. The Greene Climacteric Scale (GCS) ^19,20^, while widely used, was developed in the 1970s and lacks sexual function nuances (only 1 item) and did not define explicit thresholds to categorize overall symptom severity. The Women’s Health Questionnaire (WHQ) ^29^ includes broader symptoms but is lengthy with 36 items and was validated in a very small sample (n=48). The MENQOL ^18^, though menopause-specific, did not define symptom domains thus was not able to track women’s domain-specific changes in symptoms. The Day-to-Day Impact of Vaginal Aging (DIVA) ^37^ was developed for treatment-seeking postmenopausal women and focuses narrowly on vaginal symptoms. Collectively, these tools reflect historical strengths but are limited in their specificity, psychometric rigor, and inclusiveness of the perimenopause experience.

In summary, the Peri-SS is a concise and psychometrically robust measure of perimenopause symptom burden. By addressing the limitations of earlier instruments and offering digital deployment, it provides a modern and actionable tool for both research and clinical practice.

### Strengths and implications

In addition to strong psychometric performance, this study offers several methodological and practical strengths that support the real-world relevance and utility of the Peri-SS.

The validation process followed the COSMIN framework ^25^, ensuring rigorous evaluation across reliability, construct validity, criterion validity, and responsiveness to change. The longitudinal design enabled robust assessment of test-retest reliability and sensitivity to change—two properties often underreported in prior instruments. The sample size was adequately powered to detect differences across known groups, and the inclusion of women aged 35-59 years includes women across the reproductive stages which is broader than typically captured. While the sample was drawn from a single geographic and linguistic context (U.S.-based, English-speaking), it was younger and more racially/ethnically diverse than many existing studies, supporting the scale’s relevance to earlier and more heterogeneous perimenopause experiences.

Practically, the Peri-SS provides a digital-first, user-friendly tool that enables scalable and nuanced symptom tracking across key domains of the perimenopause experience. Its four-domain structure is clinically interpretable and supports a range of clinical and research priorities in the care of midlife women ^12^. These include evaluating and managing symptom burden, assessing the effectiveness of non-hormone and lifestyle-based interventions, monitoring the progression of symptoms such as sleep disruption and fatigue, and identifying symptom phenotypes to inform tailored support and treatment pathways ^1^.

For digital health tools, the Peri-SS enables dynamic segmentation of users based on symptom burden and supports personalization of educational content, thereby enhancing user engagement and perceived relevance ^38^. The brevity of the 16-item instrument ensures it is practical for repeated use without overwhelming users, making it especially suitable for mobile platforms and longitudinal tracking.

Furthermore, the Peri-SS has potential value beyond individual symptom monitoring. It could be deployed in workplace health initiatives or employee wellbeing platforms to assess the impact of perimenopause symptoms on productivity, absenteeism, or quality of life ^39^. This opens avenues for evaluating support interventions (e.g., policy changes, access to symptom management tools), and normalizing perimenopause and menopause-related health as a part of occupational health discourse ^40^.

By offering a validated, perimenopause-specific, and digitally deployable scale, the Peri-SS addresses a critical gap in measurement and enables more responsive, personalized, and evidence-based approaches to supporting women through perimenopause.

### Limitations and future directions

Despite its strengths, we acknowledge several limitations which would inform future research and application. First, our sample was recruited online. Although this mode of recruitment aligns with the intended context of use (digital health platforms), it nevertheless limits generalizability to individuals with lower digital literacy or limited internet access. To enhance the broader applicability of the Peri-SS, future validation studies should include populations with more diverse geographic, ethnic, educational, and digital literacy backgrounds.

Second, in this initial validation study, we relied on self-reported rather than clinically confirmed reproductive stages. However, we observed clear known-group differences in symptom burden across reproductive stages, supported by both the Peri-SS and MRS scores, which adds confidence to the validity of the self-reported categories (see Supplementary S-Tables 3 and S-Table 4). Furthermore, we conducted additional analysis on participants’ medical-seeking behaviors and found that 45.0% of participants who self-identified as perimenopausal had consulted a medical professional in the past 12 months about their symptoms. Among these, 56.8% received a formal confirmation. This finding further supports that the self-reported reproductive stage in our sample has clinical grounding. To mitigate this limitation, further research is needed to validate Peri-SS in populations with clinically confirmed reproductive stage.

Third, while the Peri-SS includes a broad range of symptoms reported by women in perimenopause, it does not capture every possible experience. Some common symptoms during perimenopause, such as headache, fatigue, and brain fog ^10^ were excluded due to poor psychometric performance (high correlation with other items or high cross domain factor loadings) during development. Additionally, menstrual cycle changes were not included as a scored symptom domain in the Peri-SS, despite being a hallmark of perimenopause ^5^. During scale development, we treated menstrual cycle irregularity as a contextual indicator for classifying women as likely perimenopausal, rather than as a symptom in its own right. However, we acknowledge that the extent and pattern of menstrual changes vary widely and may themselves be perceived by women as a meaningful part of the symptom experience. We plan to consider incorporating bleeding pattern changes as either a standalone domain or a moderating factor in future iterations, especially as more nuanced cycle-related data become available through digital tracking tools.

Finally, symptom experiences may vary across cultural and linguistic contexts. The current version of the Peri-SS was developed and validated in English among U.S.-based participants, and certain symptom concepts may not translate seamlessly. To address this, we are conducting qualitative work to adapt the scale for use in multiple languages, starting with German, French, Spanish, Brazilian Portuguese, and Italian, to ensure cultural relevance and cross-contextual validity.

## Conclusions

This study provides strong initial validation of the Peri-SS, a concise and digitally deployable self-assessment tool designed to measure the multidimensional symptom burden experienced during perimenopause. The scale demonstrated excellent psychometric properties across structural validity, internal consistency, test-retest reliability, construct validity, and responsiveness to change. Compared to legacy instruments, the Peri-SS is more inclusive of perimenopause-specific symptoms and better aligned with modern psychometric standards. Its digital-first format and clinical interpretability position it as a promising tool for research, clinical care, and digital health applications.

## Data Availability

Aggregated data analyses and deidentified individual-level data may be shared upon reasonable request for the purpose of replication, secondary analysis, or further methodological development, pending permission from Flo Health Ltd.

## Financial Disclosures / Conflicts of Interest

**YX** is a current employee of Flo Health Ltd. and holds equity interests in Flo Health Ltd.

**II** is a current employee of Flo Health Ltd. and holds equity interests in Flo Health Ltd.

**CP** is a consultant of Flo Health Ltd.

**YHM** is a current employee of Flo Health Ltd. and holds equity interests in Flo Health Ltd.

**AC** is a current employee of Flo Health Ltd. and holds equity interests in Flo Health Ltd.

**CS** is an advisor for Bayer Pharmaceutics.

**SSF** is a consultant for Era Women’s Health Platform, delivers CME lectures for PriMed, AiCME, MedAll, and Medscape, and serves on the scientific advisory board for Weight Watchers.

**LZ** is a current employee of Flo Health Ltd. and holds equity interests in Flo Health Ltd.

## Funding

No funding was received from National Institutes of Health (NIH), Wellcome Trust, or Howard Hughes Medical Institute (HHMI)

## Previous Presentations

A preprint of the manuscripts was published on medRxiv under the following DOI: https://doi.org/10.1101/2025.07.16.25331654

## Abbreviations

BMI: Body Mass Index
CFI: Comparative Fit Index
CFA: Confirmatory Factor Analysis
CI: Confidence Interval
COSMIN: COnsensus-based Standards for the selection of health Measurement Instruments
DIVA: Day-to-Day Impact of Vaginal Aging Questionnaire
EFA: Exploratory Factor Analysis
EQ-5D-5L: EuroQol five-dimensional questionnaire (five-level version)
FIML: Full-Information Maximum Likelihood
FSH: Follicle-Stimulating Hormone
GCS: Greene Climacteric Scale
HT: Hormone Therapy
ICC: Intraclass Correlation Coefficient
IRB: Institutional Review Board
MenoScale: Menopause Scale
MENQOL: Menopause-specific Quality of Life Questionnaire
MRS: Menopause Rating Scale
MTS: Menopause Transition Scale
Peri-SS: Perimenopause Symptom Scale
PROMs: Patient-Reported Outcome Measures
RMSEA: Root Mean Square Error of Approximation
SD: Standard Deviation
STROBE: Strengthening the Reporting of Observational studies in Epidemiology
SWAN: Study of Women’s Health Across the Nation
T1: Baseline Survey
T2: Follow-up Survey
WHQ: Women’s Health Questionnaire

## Supplement Digital Content

**Supplementary material A**. Self-reported menopausal status question

**Supplementary material B.** Perimenopause Symptom Scale (Peri-SS)

**Supplementary material C**. Peri-SS score categorization approach and score transformation for product display purposes

**Supplementary material D**. COSMIN reporting guideline for reporting studies of measurement properties of Patient-Reported Outcome Measures (PROMs)

**Supplementary material E**. Agreement Between MRS and Peri-SS Severity Categories

**S-Table 1**. Cross-tabulation of symptom severity categories between MRS and Peri-SS

**S-Table 2**. Item-level descriptive statistics for Peri-SS symptoms by age group among perimenopausal participants

**S-Table 3**. Descriptive statistics and known-group differences in MRS total scores by age group, menopausal status, and HT use

**S-Table 4**. Post Hoc pairwise comparisons of MRS total scores across subgroups using Wilcoxon Tests (Bonferroni-adjusted p-values)

## Notes

### Competing Interest Statement

Y.X., I.I., Y.H-M., and A.C. are employees of Flo Health Ltd. C.P. and L.Z. are contractors for Flo Health Ltd. Y.X., I.I., Y.H-M., A.C., and L.Z. hold equity interests in Flo Health Ltd. C.S. is an advisor for Bayer Pharmaceutics. S.S.F. is a consultant for Era Womens Health Platform, delivers CME lectures for PriMed, AiCME, MedAll, and Medscape, and serves on the scientific advisory board for Weight Watchers.

### Author Declarations

This study was conducted in two phases, both of which received ethical approval from an independent institutional review board. The initial scale development phase, including item generation and exploratory factor analysis, was approved by the WCB Institutional Review Board (IRB No. 20244604). The subsequent validation study, involving confirmatory factor analysis and psychometric testing, was approved under a separate protocol (IRB No. 20250554). All participants provided informed consent electronically prior to participation and were informed of their right to withdraw at any time without penalty. We compensated participants at a fair hourly rate ($12-15 USD/hour) in line with ethical guidelines for online research platforms.

### Summary of Updates

This version includes minor formatting and editorial revisions. No substantive changes were made to the study design, analyses, results, or conclusions.

