## Supplementary Materials for "Validation of the Perimenopause Symptom Scale (Peri-SS) for Digital Self-Assessment: Psychometric Evaluation Among U.S. Women Aged 35 to 59 years"

### Supplementary material A. Self-reported menopausal status question

Below is the self-report question used to classify menopausal status. Participants were presented with a standardized definition of premenopause, perimenopause, and postmenopause (with visual aid), and asked to select the stage that best described their current experience. This approach facilitated consistent classification based on cycle changes
and symptom presentation, particularly among those without a confirmed menopause date.

* 10. **How would you describe yourself:**

Please note:

- ***Perimenopause*** *is your transition through menopause. It covers the years leading up to menopause (your last ever period) and the 12 months after, at which point you can be sure you've reached menopause. Due to changing hormone levels, perimenopause is often accompanied by changes in cycle variability along with specific symptoms.*
- ***Menopause*** *is your last ever period. But you can only be sure you've reached menopause after you've gone 12 months without a period.*
- ***Premenopause*** *is the stage before perimenopause.*
- ***Postmenopause*** *is the time after your menopause. If your menopause is not confirmed yet, then you are still in perimenopause.*

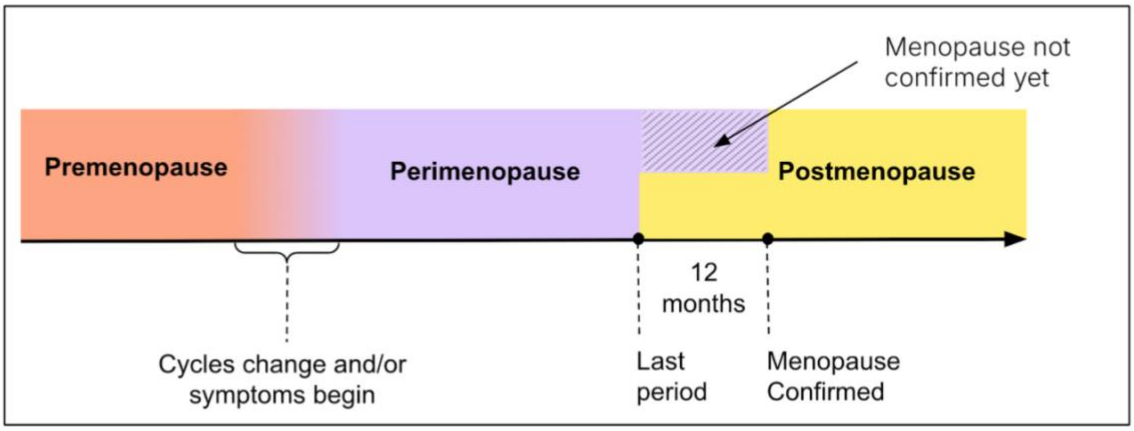


◯ I'm in perimenopause

◯ I'm premenopausal

◯ I'm postmenopausal

◯ I am unsure

◯ Prefer not to answer

### Supplementary material B. Perimenopause Symptom Scale (Peri-SS)

Below is the 16-item Perimenopause Symptom Scale (Peri-SS), a self-administered instrument designed to assess symptom burden in women undergoing perimenopause. Participants were asked to rate the severity of each symptom over the past 30 days on a 5-point Likert scale ranging from "None" to "Very Severe." The scale covers four domains: vasomotor, psychological, somatic, and sexual health symptoms.

#### Survey question

Which of the following symptoms have you experienced **in the past 30 days**? Please rate the severity of each symptom. If a symptom does not apply to you, please select "none". [Answer options: none, mild, moderate, severe, very severe]

- Hot flashes
- Night sweats
- Feeling down or sad
- Irritability
- Anxiety
- Physical/mental exhaustion
- Sleep difficulties
- Low sex drive
- Vaginal dryness
- Painful sex
- Bladder problems
- Joint/muscle pains
- Digestive issues
- Hair changes
- Skin changes
- Weight changes

**Peri-SS with explanatory note**

| Domain | Symptom item | Explanatory note |
|---|---|---|
| Sexual health | Low sex drive | low sexual desire or interest |
| | Vaginal dryness | dry or burning sensation |
| | Painful sex | Could be during or after sex |
| Psychological and emotional stress | Feeling down or sad | low mood or lack of drive |
| | Irritability | e.g. feeling nervous, tense or annoyed |
| | Anxiety | e.g. feeling on edge, restless or panicky |
| | Physical/mental exhaustion | affects memory, focus, drive and stamina |
| | Sleep difficulties | e.g. early waking or trouble falling or staying asleep / e.g. trouble falling or staying asleep |
| Vasomotor symptoms | Hot flashes | sudden heat waves in your body |
| | Night sweats | Hot flashes that happen at night |
| Physical and somatic changes | Bladder problems | peeing problems or bladder leaks |
| | Joint/muscle pains | not related to an injury or a health condition |
| | Digestive issues | e.g. bloating or constipation |
| | Hair changes | e.g. feeling dry, thin, or brittle |
| | Skin changes | e.g. acne, dryness, or itchiness |
| | Weight changes | e.g. weight loss or gain |

### Supplementary material C. Peri-SS score categorization approach, calculation process, and score transformation for product display purposes

To support interpretation, we categorise Peri-SS into four severity categories, conceptually aligned with the MRS. The severity categories were empirically derived using a multinomial logistic regression model, with MRS category as the outcome and the Peri-SS score, EQ-5D utility, and age as predictors. Predicted probabilities for each MRS category were plotted across the Peri-SS score range, and thresholds were identified at the cross-over points where one severity category became more probable than the next. This approach allowed for a data-driven calibration of symptom burden categories while accounting for health-related quality of life and age. Based on this analysis, Peri-SS score thresholds were defined as follows: 0–8 (none to mild), 9–18 (mild to moderate), 19–35 (moderate to severe), and 36 or above (severe).

The Peri-SS total score is calculated using an equal-weighted by domain approach rather than by simply summing responses across the 16 individual items.

Items are grouped into four domains: vasomotor symptoms (2 items), psychological and emotional symptoms (5 items), sexual health symptoms (3 items), and physical and somatic symptoms (6 items). And each item is scored from 0 (“none”) to 4 (“very severe”).

For each participant, we first calculate the mean score within each of the four domains. We then take the average of those four domain scores and scale this to a 0–100 range:

$$\text{Peri-SS score} = \frac{D_{vasomotor} + D_{psychological} + D_{sexual} + D_{somatic}}{4} \times 25$$

For example, suppose a participant gives the following responses to the 16 items, we can calculate the domain score by taking the average score within each domain.

| Domain | Item scores | Domain score |
|---|---|---|
| Vasomotor, 2 items | 1, 2 | 1.50 |
| Psychological/emotional, 5 items | 2, 2, 1, 3, 2 | 2.00 |
| Sexual health, 3 items | 1, 1, 0 | 0.67 |
| Physical/somatic, 6 items | 2, 2, 1, 1, 2, 2 | 1.67 |

Then the total Peri-SS score can be calculated as follows:

$$\text{Peri-SS score} = \frac{1.50 + 2.00 + 0.67 + 1.67}{4} \times 25 = 36.5$$

In digital product contexts, we also developed a non-linear transformation of the Peri-SS score to improve interpretability in user-facing digital applications. This transformation enhances interpretability for end users and reduces the visual overemphasis of mid-range values. It is intended solely for display purposes within the product interface and is not used in clinical or research analyses.

This transformation compresses the upper range of the scale, making the range aligns better with symptom severity as perceived in user testing.

The raw Peri-SS score is first calculated as the average of four domain scores (each scaled to 0–100). The final product-facing score is then derived using the following transformation:

$$Final\ PeriSS\ score = 10\ \times \sqrt{Raw\ PeriSS\ score}$$

This transformation is intended solely for display within the product interface and is not used for clinical analysis or psychometric validation. It was developed in collaboration with the product and user experience teams at Flo Health Limited to ensure that moderate scores (e.g., 30–40) are perceived as more intuitive and actionable by users.

**Supplementary material D. COSMIN reporting guideline for reporting studies of measurement properties of Patient-Reported Outcome Measures (PROMs)**

| Item | Item name | Guidance for reporting | Reported in section |
|---|---|---|---|
| **Report section: Title** | | | |
| T1 | Title | Indicate that the study focuses on the measurement properties of a PROM. | Title |
| **Report section: Abstract** | | | |
| A1 | Objectives | Provide the specific objective(s) of the research, specifying (1) the name (and version, if relevant), and construct(s) of the PROM, (2) the measurement properties being evaluated, and (3) relevant study characteristics. | Abstract |
| A2 | Design | Specify (details of the) study design used to evaluate the measurement properties. | Abstract |
| A3 | Methods | Specify the methods for evaluating each measurement property. | Abstract |
| A4 | Results | Provide the main results for all measurement properties evaluated. | Abstract |
| A5 | Discussion/ Conclusion | Provide a brief statement of the implications of the findings in the context of existing evidence on the PROM. | Abstract |
| **Report section: Introduction** | | | |
| GM1 | Study design | Specify (details of the) study design used to evaluate the measurement properties. | Methods – Scale development and validation study design |
| GM2 | Participants | Specify how the study participants were selected. Specify the inclusion and exclusion criteria | Methods – Participants and recruitment |
| GM3 | PROM details | Provide details about the original version of the PROM as well as of the PROM version being studied, specify the conceptual framework (reflective/formative model), details on the structure (the number of items and subscales), the language, response scale, recall period, direction of scoring, and scoring algorithm of the PROM. Specify how the PROM was administered (e.g., in what setting, mode of administration (e.g. paper, electronic) what instructions were given), including the country in which it is administered | Methods – The Perimenopause Symptom Scale (Peri-SS) |

| | | | |
|---|---|---|---|
| GM4 | Additional data collection | Describe why and how other data was collected (e.g., construct and measurement properties of the comparator instruments, characteristics of groups being compared, and rationale for choosing groups), including mode of administration (e.g., paper, electronic). | Methods – Validation procedures |
| GM5 | Time points procedures | Provide all time points of all measurements. | Methods – Participants and recruitment / Validation procedures |
| GM6 | Justification for sample size | Provide a rationale for the sample size for all measurement properties analyses (including subgroups). | Methods – Scale development and validation study design |
| GM7 | Statistical analyses | Describe the statistical analyses corresponding to all objectives (see measurement properties specific boxes). Describe the criteria for good measurement properties. Name the statistical package used and the version. | Methods – Validation procedures |
| GM8 | Missing data | Describe approaches for dealing with missing data. | Methods – Validation procedures; Figure 1 |
| GM9 | Unplanned analysis | Specify analyses that were unplanned and their rationale. | Not applicable |
| **Report section: General results** | | | |
| GR1 | Participant characteristics | Provide study participants' characteristics, specified per subgroup if applicable. | Results – Participant characteristics, Table 1 |
| GR2 | Sample size | Provide the total number of participants included in the study and the sample size for each analysis. | Results – Participant characteristics, Figure 1 |
| GR3 | Missing data | Provide amount of (proportion or count) and reasons for missing data for each analysis for the PROM, and for any | Results – Figure 1 |

| | | | |
|---|---|---|---|
| | | analyses of other outcome measurement instruments. | |
| GR4 | Results | Describe the results corresponding to all objectives (see measurement properties specific boxes). | Results – All subsections |
| **Report section: Discussions/conclusions** | | | |
| DC1 | Measurement property evidence | Provide the main findings and if each measurement property is sufficient or insufficient and why. | Discussion – Principal findings |
| DC2 | Practical relevance | Discuss the practical relevance of the findings in terms of recommendations for (not) using the PROM. | Discussion – Strengths and implications |
| DC3 | Strengths and limitations | Discuss strengths and limitations of each study. For example, discuss if there were any potential biases in the study that could have impacted the results. | Discussion – Strengths and implications; Limitations |
| DC4 | Generalizability | Discuss generalizability of the results. For example, discuss whether the results could be generalized to other populations given the sample studied. | Discussion – Limitations and future directions |
| DC5 | Instrument changes | Discuss what modifications are needed to the existing PROM. | Discussion – Limitations and future directions |
| DC6 | Future research | Describe new research questions or hypotheses generated from these findings, and provide/describe the research needed to answer those questions. | Discussion – Limitations and future directions |
| DC7 | Conclusions | Provide the overall conclusions for the use of the PROM. | Discussion – Final paragraph |
| GM1 | Study design | Specify (details of the) study design used to evaluate the measurement properties. | Methods – Scale development and validation study design |
| GM2 | Participants | Specify how the study participants were selected. Specify the inclusion and exclusion criteria | Methods – Participants and recruitment |
| GM3 | PROM details | Provide details about the original version of the PROM as well as of the PROM version being studied, specify the conceptual framework (reflective/formative model), details on the structure (the number of items and subscales), the language, response scale, recall period, direction of scoring, and scoring algorithm of the PROM. Specify how the PROM was administered (e.g., in what setting, mode of administration (e.g. paper, electronic) what | Methods – The Perimenopause Symptom Scale (Peri-SS) |

| | | | |
|---|---|---|---|
| | | instructions were given), including the country in which it is administered | |
| GM4 | Additional data collection | Describe why and how other data was collected (e.g., construct and measurement properties of the comparator instruments, characteristics of groups being compared, and rationale for choosing groups), including mode of administration (e.g., paper, electronic). | Methods – Validation procedures |
| GM5 | Time points procedures | Provide all time points of all measurements. | Methods – Participants and recruitment / Validation procedures |
| **Report section: Other information** | | | |
| O1 | Conflict of interest | State any conflict of interest you may have related to the PROM. This may include any involvement in the development of the PROM or any commercial funding or profit. | Statements and declarations - conflict of interests |

**Supplementary material E. Agreement Between MRS and Peri-SS Severity Categories**

To explore the extent to which Peri-SS categorization aligned with an established measure, we assessed categorical agreement between the Peri-SS and MRS severity levels (minimal, mild, moderate, severe) using Cohen's kappa. A total of $n$ = 633 participants who self-identified as in perimenopause were included in the analysis. The unweighted kappa coefficient was 0.58 (95% CI: 0.53–0.63), indicating moderate agreement between the two scales. S-Table 1 presents the detailed categorical agreement between MRS and Peri-SS severity levels. This level of agreement suggests Peri-SS captures symptom severity in a way that is broadly consistent with the MRS, but also potentially captures additional or slightly different dimensions, consistent with our scale development goals.

**S-Table 1.** Cross-tabulation of symptom severity categories between MRS and Peri-SS

| | | Severity level by MRS | | | |
|---|---|---|---|---|---|
| | | **Mild** | **Minimal** | **Moderate** | **Severe** |
| **Symptom severity level by Peri-SS** | **Mild** | **47** | 26 | 19 | 0 |
| | **Minimal** | 2 | **26** | 1 | 0 |
| | **Moderate** | 47 | 8 | **196** | 27 |
| | **Severe** | 5 | 1 | 43 | **185** |

**S-Table 2.** Item-level descriptive statistics for Peri-SS symptoms by age group among perimenopausal participants

| **Domain** | **Item** | **35-44 (N=184)** | **45-54 (N=435)** | **55-59 (N=14)** | **Overall (N=633)** |
|---|---|---|---|---|---|
| **Vasomotor symptoms** | Hot flashes | 0.91 (0.93) | 1.20 (1.00) | 1.40 (1.20) | 1.10 (1.00) |
| | Night sweats | 1.40 (1.00) | 1.30 (1.10) | 1.30 (1.10) | 1.30 (1.00) |
| **Psychological and emotional stress** | Feeling down or sad | 1.50 (0.98) | 1.60 (1.00) | 0.86 (0.95) | 1.50 (0.99) |
| | Irritability | 1.70 (0.98) | 1.70 (0.97) | 1.20 (1.10) | 1.70 (0.98) |
| | Anxiety | 1.70 (0.99) | 1.70 (1.10) | 0.93 (1.00) | 1.70 (1.10) |
| | Physical/mental exhaustion | 1.80 (1.10) | 1.80 (1.10) | 1.30 (0.99) | 1.80 (1.10) |
| | Sleep difficulties | 1.70 (1.10) | 1.90 (1.10) | 1.90 (1.20) | 1.80 (1.10) |
| **Sexual health** | Low sex drive | 1.50 (1.30) | 1.80 (1.30) | 1.80 (1.50) | 1.70 (1.30) |
| | Vaginal dryness | 0.95 (1.10) | 1.00 (1.10) | 1.10 (1.20) | 1.00 (1.10) |
| | Painful sex | 0.57 (0.92) | 0.61 (1.00) | 0.93 (1.10) | 0.61 (0.98) |
| **Physical and somatic changes** | Bladder problems | 0.76 (0.98) | 0.86 (0.99) | 1.50 (1.30) | 0.85 (1.00) |
| | Joint/muscle pains | 1.30 (1.00) | 1.50 (1.10) | 1.80 (1.30) | 1.50 (1.10) |
| | Digestive issues | 1.10 (1.10) | 1.20 (1.00) | 1.10 (1.00) | 1.10 (1.00) |
| | Hair changes | 1.20 (1.20) | 1.20 (1.10) | 0.86 (1.20) | 1.20 (1.10) |
| | Skin changes | 1.00 (1.00) | 1.10 (0.99) | 0.71 (0.99) | 1.10 (1.00) |
| | Weight changes | 1.20 (1.00) | 1.40 (1.10) | 1.40 (1.20) | 1.30 (1.10) |

*Note:* Values are presented as Mean (SD). Items are grouped by symptom domain.

**S-Table 3.** Descriptive statistics and known-group differences in MRS total scores by age group, menopausal status, and HT use

| Group Variable | Category | N | Mean (SD) | Median [Min, Max] | Test statistic | p-value |
|---|---|---|---|---|---|---|
| Overall | | 1255 | 12.6 (7.48) | 12.0 [0, 39.0] | | |
| Age group | 35–44 | 497 | 11.5 (7.38) | 11.0 [0, 38.0] | $\chi^2$ = 20.0, df = 2 | <0.001 |
| | 45–54 | 691 | 13.4 (7.47) | 13.0 [0, 39.0] | | |
| | 55–59 | 67 | 12.4 (7.30) | 12.0 [0, 32.0] | | |
| Menopausal status | Premenopause | 327 | 10.7 (7.21) | 10.0 [0, 37.0] | $\chi^2$ = 43.2, df = 3 | <0.001 |
| | Perimenopause | 633 | 13.8 (7.43) | 13.0 [0, 38.0] | | |
| | Postmenopause | 119 | 13.5 (7.93) | 13.0 [0, 39.0] | | |
| | Unsure | 176 | 11.3 (7.01) | 10.0 [0, 33.0] | | |
| HT use | No | 1190 | 12.5 (7.49) | 12.0 [0, 39.0] | $\chi^2$ = 7.02, df = 1 | 0.008 |
| | Yes | 65 | 14.9 (7.07) | 13.0 [1.00, 34.0] | | |

**S-Table 4.** Post Hoc pairwise comparisons of MRS total scores across subgroups using Wilcoxon Tests (Bonferroni-adjusted p-values)

| Group | Category | Test statistic | p-value | p-value adjusted |
|---|---|---|---|---|
| Age group | 35–44 vs 45–54 | w = 145,614 | <0.001 | <0.001 |
| | 35–44 vs 55–59 | w = 15,221 | 0.254 | 0.508 |
| | 45–54 vs 55–59 | w = 24,630 | 0.386 | 0.508 |
| Menopausal status | Peri vs Pre | w = 38,526 | 0.692 | 0.692 |
| | Peri vs Post | w = 128,168 | <0.001 | <0.001 |
| | Peri vs Unsure | w = 66,072 | <0.001 | <0.001 |
| | Post vs Pre | w = 23,512 | <0.001 | 0.003 |
| HT use | Post vs Unsure | w = 12,112 | 0.022 | 0.067 |
| | Pre vs Unsure | w = 27,136 | 0.291 | 0.582 |
| | HT use vs No | w = 31,143 | 0.008 | 0.008 |
